# The Influence of Age, Sex, and Socioeconomic Status on Glycemic Control Among People with Type 1 and Type 2 Diabetes in Canada: A Patient-Led Longitudinal Retrospective Cohort Study

**DOI:** 10.1101/2021.12.14.21267759

**Authors:** Seyedmostafa Mousavi, Dana Greenberg, Ruth Ndjaboué, Michelle Greiver, Olivia Drescher, Selma Chipenda Dansokho, Denis Boutin, Jean-Marc Chouinard, Sylvie Dostie, Robert Fenton, Marley Greenberg, Jon McGavock, Adhiyat Najam, Monia Rekik, Tom Weisz, Donald J. Willison, Audrey Durand, Holly O. Witteman

**Affiliations:** Universite Laval; Diabetes Action Canada; Universite de Sherbrooke; University of Toronto; University of Manitoba

## Abstract

**Background:** Clinical guidelines for most adults with diabetes recommend maintaining hemoglobin A1c (HbA1c) ≤7.0% (<53 mmol/mol) to avoid microvascular and macrovascular complications. People with diabetes of different ages, sexes, and socioeconomic statuses may differ in their ease of attaining this goal. As a team of people with diabetes, researchers, and health professionals, we aimed to explore patterns in HbA1c results among people with type 1 or type 2 diabetes in Canada. Our research question was identified by people living with diabetes.

**Methods:** We used generalized estimating equations to analyze the effects of age, sex and socioeconomic status in 947,543 HbA1c results measured from 2010 to 2019 among 90,770 people living with type 1 or 2 diabetes in Canada. People living with diabetes reviewed and interpreted the results.

**Results:** HbA1c results at or below 7.0% represented 30.5% (male people living with type 1 diabetes), 21.0% (female people living with type 1 diabetes), 55.0% (male people living with type 2 diabetes) and 59.0% (female people living with type 2 diabetes) of results in each subcategory. We observed higher HbA1c values during adolescence and, for people living with type 2 diabetes, among people living in lower income areas. Among those with type 1 diabetes, female people tended to have lower HbA1c than male people during childbearing years but higher HbA1c than male people during menopausal years. Team members living with diabetes confirmed that the patterns we observed reflected their own life courses and suggested these results be communicated to health professionals and other stakeholders to improve treatment for people living with diabetes.

**Interpretation:** A substantial proportion of people with diabetes in Canada are insufficiently supported to maintain guideline-recommended glycemic control goals. Blood sugar management goals may be particularly challenging for people who are going through adolescence, menopause, or living with fewer financial resources. Health professionals should be aware of the challenging nature of glycemic management and policymakers in Canada should provide more support for people with diabetes to live healthy lives.

## Introduction

Diabetes is a chronic condition with two common types: type 1 and type 2. [1] Both types of diabetes are marked by elevated blood glucose levels but their causes of onset and treatments are generally different [2] and type 1 is less common than type 2. [3] A standard laboratory test for people with all types of diabetes is a hemoglobin A1c or HbA1c test. This test measures the concentration in a person’s blood of a compound called glycated hemoglobin, which is created when glucose molecules combine with hemoglobin in the blood. Unlike a single blood glucose measure, HbA1c is a measure of approximate mean blood glucose over 2-3 months [4] that can be measured at any time of day [5] and is often used as a marker of overall glycemic control. Higher HbA1c is associated with increased prevalence of complications of diabetes, affecting eyes, kidneys, heart, and nerves. [1] Clinical guidelines in Canada recommend maintaining HbA1c ≤7.0% (≤53 mmol/mol) for most adults with diabetes and ≤7.5% (≤59 mmol/mol) for most children with diabetes. [6]

Among people with diabetes, differences in HbA1c are associated with sociodemographic characteristics, including age [7–10], socioeconomic status [11,12], and sex. [9,13–23] Specifically, HbA1c tends to be higher among adolescents compared to other age groups [7–10] and lower among people with higher socioeconomic status (measured using postal codes) compared to those with lower socioeconomic status. [11,12] Studies in comparing HbA1c by sex have shown mixed results across countries, sometimes showing higher HbA1c among those who are male [9], sometimes higher HbA1c among those who are female [13–20] and other times no difference. [21–23]

Relationships between HbA1c and individual and social characteristics are not only apparent in the literature [24]; they are also tangible in the lives of people with diabetes. Many people living with diabetes are aware—through reflections on their own lives and comparisons with peers— that HbA1c goals may be easier to attain in some situations than others. Such expertise gleaned from lived experience of diabetes can add insight and nuance to diabetes research. Including this expertise in health research is a central tenet of patient partnership. Patient-partnered research involves people living with conditions as full members of the research team. In addition to moral reasons that people living with conditions should be included in research decisions that affect them, such inclusion can also improve studies’ relevance and quality. [25– 27]

This study’s research question was developed by people living with type 1 or type 2 diabetes involved in a larger project in which people living with diabetes developed and prioritized research questions about diabetes. The aim of this study was to answer one of the identified and prioritized questions: ‘Accounting for socioeconomic status, what patterns exist in HbA1c results among people in Canada with type 1 or type 2 diabetes of different sexes at different ages?’

## Methods

### Study design

The study used a longitudinal retrospective cohort design.

### Study Data and Inclusion Criteria

To analyze our research question, we used data from the Canadian National Diabetes Repository. This repository consists of electronic medical records for approximately 123,543 (as of July 1, 2021) people with type 1 and 2 diabetes living in Canada. The data are thus far collected from patients treated in family medicine practices from 5 Canadian provinces: Ontario, Alberta, Manitoba, Quebec, and Newfoundland & Labrador respectively. [28,29] Medical records in Canada do not currently specify type of diabetes. We therefore distinguished people with type 1 and type 2 diabetes using an algorithm recently developed and validated with Canadian data. The machine learning algorithm analyzes 21 variables (e.g., insulin use, non-metformin antihyperglycemic agent use, insulin pump use) and demonstrated sensitivity (i.e., ability to correctly identify that someone with type 1 diabetes has type 1 diabetes) of 80.6% and specificity (i.e., ability to correctly identify that someone without type 1 diabetes does not have type 1 diabetes) of 99.8%. [30]

For this study, we included all HbA1c results in the the Canadian National Diabetes Repository from everyone with diabetes who had at least one HbA1c measurement between 2010 and 2019 and for whom our other variables of interest (age, sex, socioeconomic status) were available. In other words, we excluded any records lacking one or more of these data elements. We also excluded records with HbA1c results under 3.5% (15 mmol/mol) or above 20% (195 mmol/mol) as clinical experts on our team deemed these to be likely laboratory errors, data entry errors, or rare outliers. We derived individuals’ ages by subtracting each person’s date of birth from the date when each HbA1c measurement was performed. We excluded data from individuals under age 10, as they contributed such a small proportion of the data in the repository (0.04%) that we would be unable to conduct robust analyses for this subpopulation. As a proxy measure of socioeconomic status, the Canadian National Diabetes Repository applies an established index developed by the Canadian Institute for Health Information [31] to derive neighbourhood before-tax income quintiles using the first three digits of individuals’ most recent residential postal code. Quintile 1 denotes people living in lower-income neighborhoods, quintiles 2-4 denote middle-income neighborhoods, and quintile 5 is assigned to highest-income neighborhoods. [32] Our study received ethics approval from the Laval University Research Ethics Committee, 2020-373/ 20-01-2021.

### Statistical Analysis

Our unit of analysis was each HbA1c result. We inspected HbA1c results graphically across ages using loess curves for type 1 diabetes. We used gam curves for type 2 diabetes to account for the large amount of data in this category. We then analyzed HbA1c results using Generalized Estimating Equations to account for dependency structure in between-subject and within-subject levels. As the correlations between HbA1c measurements in different years were similar, we used an exchangeable correlation structure to account for correlations between HbA1c measurements of each individual. [33] Because the distribution of HbA1c was highly skewed, we used log transformation to reduce the skewness and stabilize the variance. [34] Due to nonlinearity, we categorized age, dividing it into ordinal categories at 10 or 20-year intervals. Age range 10-19 includes puberty and menarche for a large proportion of people who experience puberty and menarche. [35,36] Age range 20-39 includes childbearing years for a large proportion of people who experience pregnancy. [37–39] Age range 40-59 includes perimenopause & menopause for a large proportion of people who experience these. [40–43] Age range 60-79 includes postmenopause for a large proportion of people who experience postmenopause. [44] Age range 80 and older includes advanced adulthood.

We performed two-way analysis of variance (ANOVA) to determine the relationship between HbA1c and independent variables. We then analyzed potential interactions between age and sex for type 1 and 2 diabetes, reporting pairwise interaction contrasts. We set our threshold for statistical significance at 0.05. We performed all analyses using R 3.6.1 and associated packages, including ggplot2 for graphics, GEE for Generalized Estimating Equations, and Estimated Marginal Means for contrasts. [45–55]

### Interpreting results with team members with diabetes

Following our analyses, we organized meetings with members of the research team living with type 1 and type 2 diabetes. These team members had already been involved throughout the year-long larger project, including a series of early meetings to orient all team members to epidemiological cohort studies, and subcommittee meetings led and attended only by patient partners to generate research questions.

To present and discuss results of this study, our bilingual team held one meeting in English and one meeting in French. We invited all team members living with diabetes to attend, presented the results for about 15 minutes, and then held an open discussion for the remaining 45 minutes. We recorded meetings to ensure we accurately noted team members’ comments and invited all team members to review the manuscript to ensure we conveyed meaning correctly. We invited all team members to fulfil authorship criteria as defined by the International Committee of Medical Journal Editors and to accordingly be co-authors on the manuscript. Nine team members accepted the invitation to serve as manuscript co-authors.

## Results

### Population

The resulting dataset consisted of two groups of data: one for people identified by the algorithm as having type 1 diabetes, one for people identified by the algorithm as having type 2 diabetes. Table 1 summarizes HbA1c results from people with type 1 and 2 diabetes by age, sex and socioeconomic status. Because HbA1c continues to be reported as a percentage in Canada, we use percentages to facilitate understanding by people living with diabetes in Canada. Out of 1950 and 946,931 available records, we excluded 1 record from the data available for people with type 1 diabetes and 1337 records from the data available for people with type 2 diabetes due to HbA1c values under 3.5% (15 mmol/mol) or above 20% (195 mmol/mol). Mean HbA1c was 8.3% (67 mmol/mol) for female people with type 1 diabetes, 8.0% (64 mmol/mol) for male people with type 1 diabetes, 7.1% (54 mmol/mol) for female people with type 2 diabetes, and 7.2% (55 mmol/mol) for male people with type 2 diabetes. In both types of diabetes, people who are female more often lived in geographic areas with lower socioeconomic status compared to people who are male.

**Table 1.**
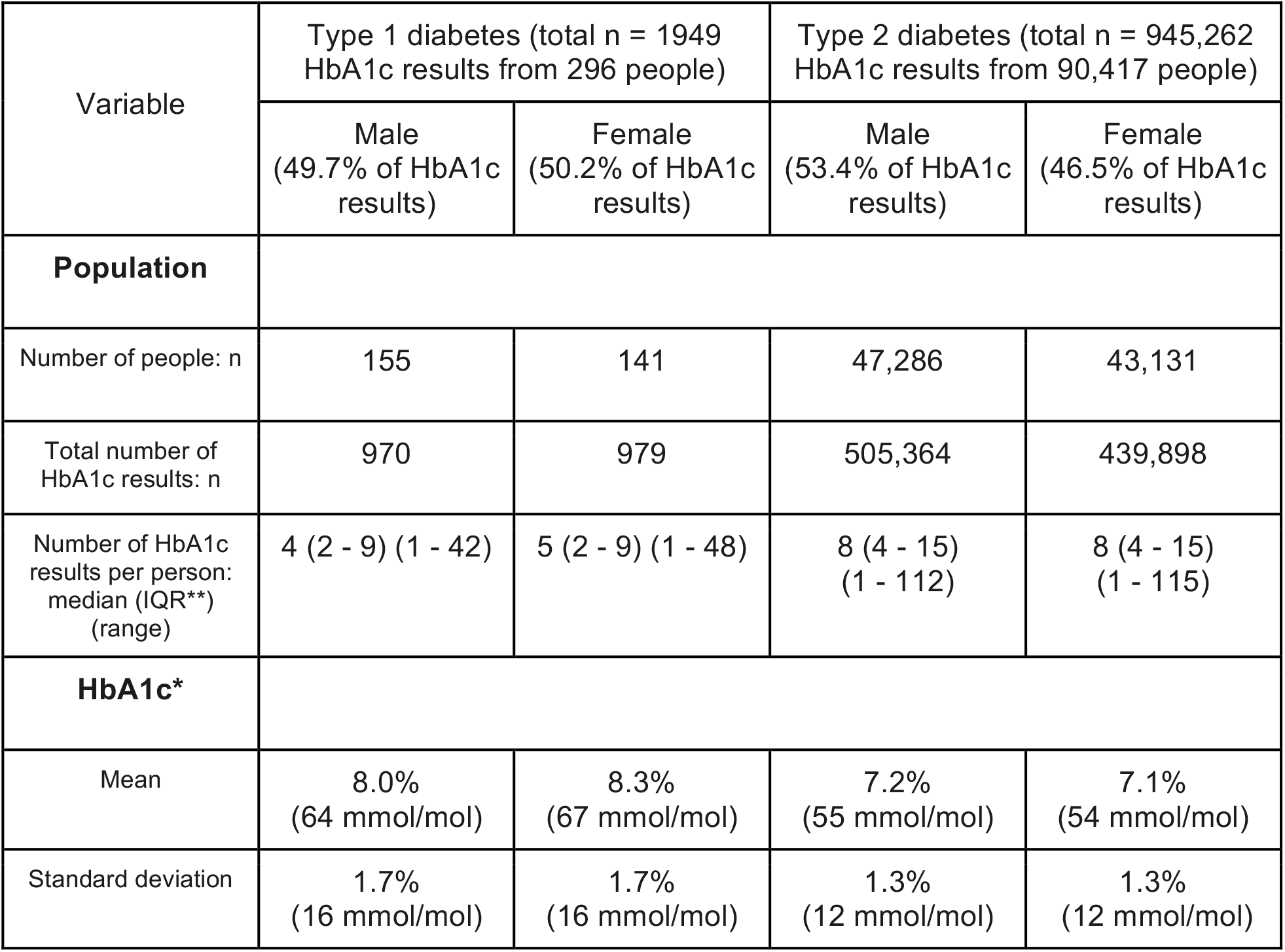
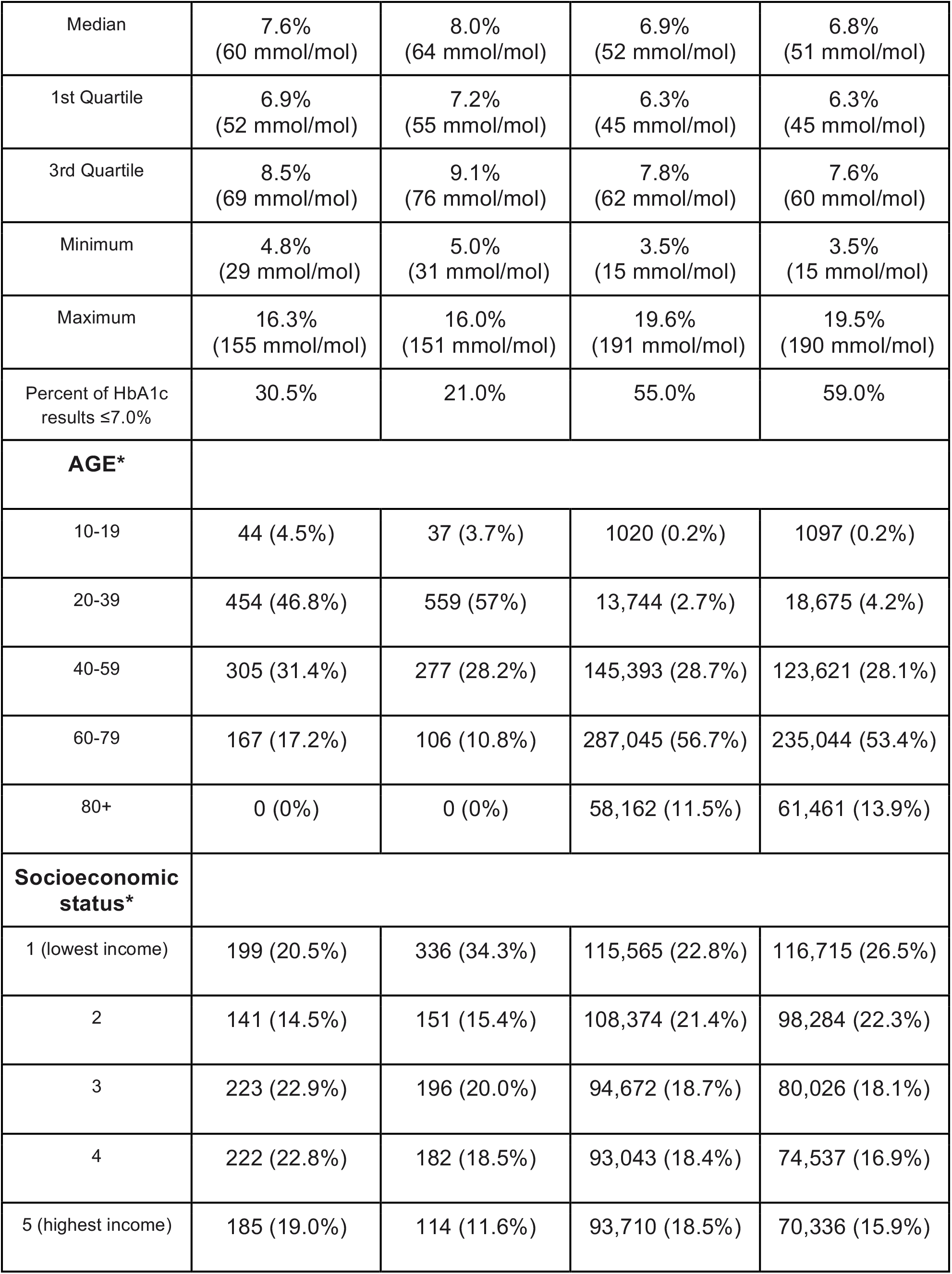
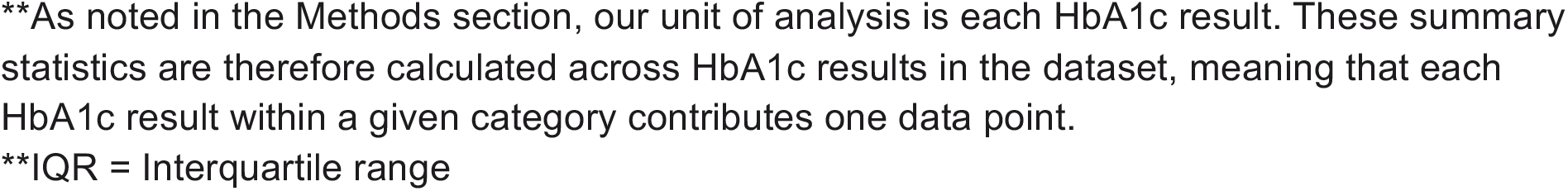
Characteristics of HbA1c results* among people with type 1 and 2 diabetes.

| Variable | Type 1 diabetes (total n = 1949<br>HbA1c results from 296 people) |  | Type 2 diabetes (total n = 945,262<br>HbA1c results from 90,417 people) |  |
| --- | --- | --- | --- | --- |
|  | Male<br>(49.7% of HbA1c<br>results) | Female<br>(50.2% of HbA1c<br>results) | Male<br>(53.4% of HbA1c<br>results) | Female<br>(46.5% of HbA1c<br>results) |
| **Population** |  |  |  |  |
| Number of people: n | 155 | 141 | 47,286 | 43,131 |
| Total number of<br>HbA1c results: n | 970 | 979 | 505,364 | 439,898 |
| Number of HbA1c<br>results per person:<br>median (IQR**) (range) | 4 (2 - 9) (1 - 42) | 5 (2 - 9) (1 - 48) | 8 (4 - 15)<br>(1 - 112) | 8 (4 - 15)<br>(1 - 115) |
| **HbA1c\*** |  |  |  |  |
| Mean | 8.0%<br>(64 mmol/mol) | 8.3%<br>(67 mmol/mol) | 7.2%<br>(55 mmol/mol) | 7.1%<br>(54 mmol/mol) |
| Standard deviation | 1.7%<br>(16 mmol/mol) | 1.7%<br>(16 mmol/mol) | 1.3%<br>(12 mmol/mol) | 1.3%<br>(12 mmol/mol) |
| Median | 7.6%<br>(60 mmol/mol) | 8.0%<br>(64 mmol/mol) | 6.9%<br>(52 mmol/mol) | 6.8%<br>(51 mmol/mol) |
| 1st Quartile | 6.9%<br>(52 mmol/mol) | 7.2%<br>(55 mmol/mol) | 6.3%<br>(45 mmol/mol) | 6.3%<br>(45 mmol/mol) |
| 3rd Quartile | 8.5%<br>(69 mmol/mol) | 9.1%<br>(76 mmol/mol) | 7.8%<br>(62 mmol/mol) | 7.6%<br>(60 mmol/mol) |
| Minimum | 4.8%<br>(29 mmol/mol) | 5.0%<br>(31 mmol/mol) | 3.5%<br>(15 mmol/mol) | 3.5%<br>(15 mmol/mol) |
| Maximum | 16.3%<br>(155 mmol/mol) | 16.0%<br>(151 mmol/mol) | 19.6%<br>(191 mmol/mol) | 19.5%<br>(190 mmol/mol) |
| Percent of HbA1c<br>results $\leq 7.0\%$ | 30.5% | 21.0% | 55.0% | 59.0% |
| **AGE\*** |  |  |  |  |
| 10-19 | 44 (4.5%) | 37 (3.7%) | 1020 (0.2%) | 1097 (0.2%) |
| 20-39 | 454 (46.8%) | 559 (57%) | 13,744 (2.7%) | 18,675 (4.2%) |
| 40-59 | 305 (31.4%) | 277 (28.2%) | 145,393 (28.7%) | 123,621 (28.1%) |
| 60-79 | 167 (17.2%) | 106 (10.8%) | 287,045 (56.7%) | 235,044 (53.4%) |
| 80+ | 0 (0%) | 0 (0%) | 58,162 (11.5%) | 61,461 (13.9%) |
| **Socioeconomic**<br>**status\*** |  |  |  |  |
| 1 (lowest income) | 199 (20.5%) | 336 (34.3%) | 115,565 (22.8%) | 116,715 (26.5%) |
| 2 | 141 (14.5%) | 151 (15.4%) | 108,374 (21.4%) | 98,284 (22.3%) |
| 3 | 223 (22.9%) | 196 (20.0%) | 94,672 (18.7%) | 80,026 (18.1%) |
| 4 | 222 (22.8%) | 182 (18.5%) | 93,043 (18.4%) | 74,537 (16.9%) |
| 5 (highest income) | 185 (19.0%) | 114 (11.6%) | 93,710 (18.5%) | 70,336 (15.9%) |

\*\*As noted in the Methods section, our unit of analysis is each HbA1c result. These summary statistics are therefore calculated across HbA1c results in the dataset, meaning that each HbA1c result within a given category contributes one data point.
\*\*IQR = Interquartile range

As shown in Figure 1, we observed potential relationships between age and HbA1c among people with type 1 or type 2 diabetes. We also observed that such relationships may differ between those who are male and female. As shown in Figure 2, socioeconomic status may also be associated with HbA1c for people with type 1 and type 2 diabetes during much of adulthood, with higher HbA1c values among adults aged 30 or 40 through 70 and living in geographic areas with lower mean income.

**Figure 1.**
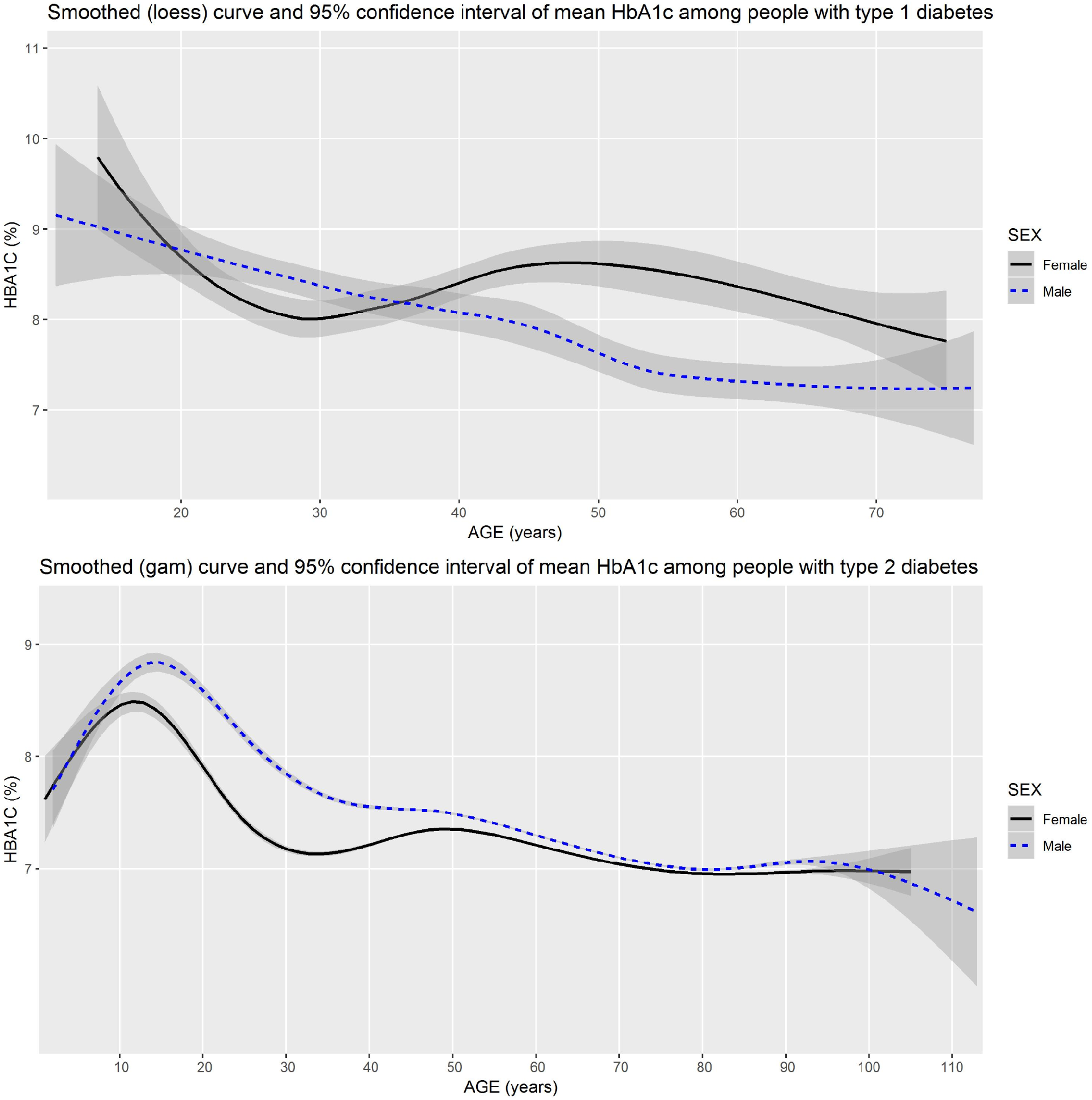
Mean HbA1c of male and female people across all ages of people with type 1 and type 2 diabetes in Canada.

**Figure 2.**
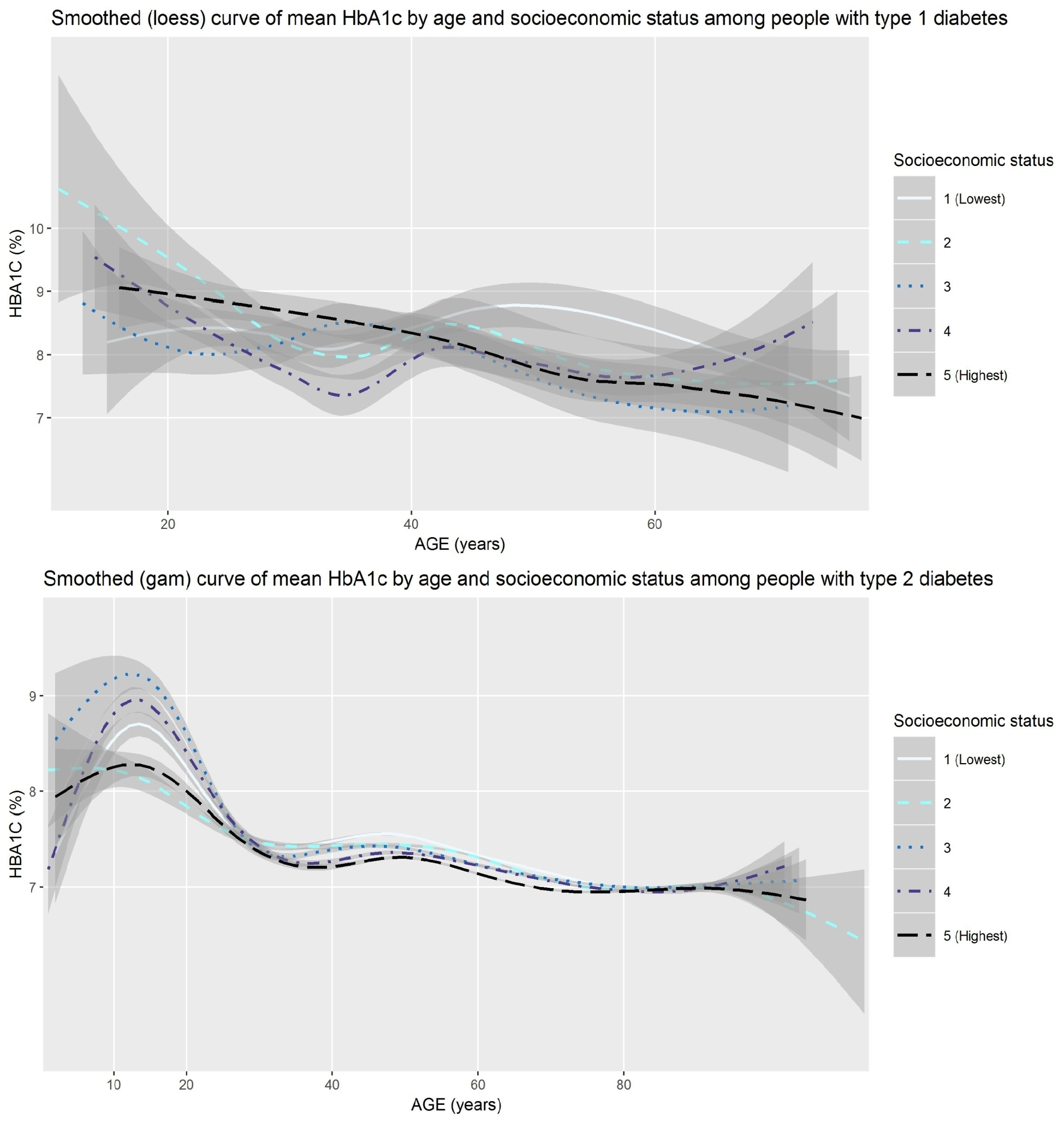
Mean HbA1c of people with type 1 and type 2 diabetes in Canada across all ages by socioeconomic status.

Table 2 shows the results of the ANOVA fitted by generalized estimating equations for both types of diabetes.

**Table 2.**
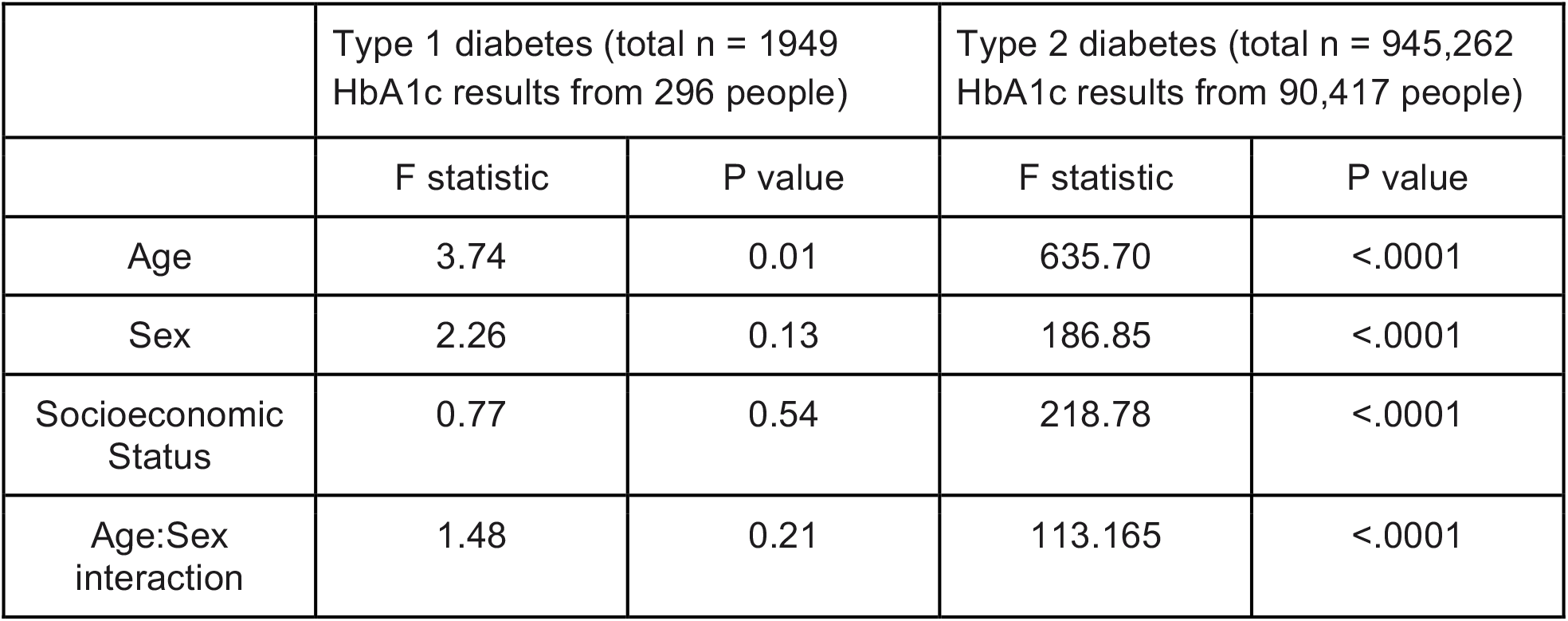
Relationships between age, sex, socioeconomic status and HbA1c among people with type 1 and type 2 diabetes in Canada.

According to our analyses of 296 people in the Canadian National Diabetes Repository who have type 1 diabetes, there is a statistically significant relationship between age and HbA1c, with overall lower HbA1c among older people with type 1 diabetes. We observed no statistically significant relationships between sex and HbA1c, socioeconomic status and HbA1c, and the interaction term showed no statistically significant differences between people who are male and female at different ages. For 90,417 people in the Canadian National Diabetes Repository who have type 2 diabetes, all variables demonstrate statistically significant relationships with HbA1c. HbA1c results are lower among people who are older, higher among people who are male, and higher among people living in geographic areas with lower mean income. There is a significant interaction between age and sex. Supplementary materials show pairwise interaction contrasts for this interaction.

### Interpretation of results by team members with diabetes

Approximately equal numbers of team members living with type 1 and type 2 diabetes attended the meetings to discuss and interpret results. In these meetings, people living with diabetes raised potential post hoc explanations for the findings, asked technical questions about the analyses, raised potential study limitations, and discussed implications for policy and future research.

Specifically, with respect to explanations of findings, women living with type 1 diabetes suggested that the potential pattern among female people with type 1 diabetes mimicked their own life courses and may reflect lower HbA1c during potential childbearing years and higher HbA1c during potential menopausal years. People with diabetes also noted the differing sex-based differences between type 1 diabetes and type 2 diabetes. Although in both types of diabetes, HbA1c results for female people dipped around age 30 and peaked just before age 50 while HbA1c for male people descended more smoothly with age, in the case of type 1 diabetes, curves met and intersected at multiple ages whereas there were no similar meetings and intersections in the case of type 2 diabetes. Finally, people with diabetes questioned whether differences in HbA1c at different ages might reflect differences in the effort that people are able to put into management at different stages of the life course, depending on when they were diagnosed.

With respect to technical questions, people living with diabetes noted that accurate HbA1c measurement is not possible for some individuals [56], queried how hypoglycemia unawareness might influence HbA1c results, and raised the issue that HbA1c is a highly imperfect measure. HbA1c is essentially analogous to average blood glucose, and averages can mask substantial variation. Nonetheless, it remains a standard measure, as other methods of measurement (e.g., time-in-range measured by flash or continuous glucose monitoring) are not universally available across Canada.

People living with diabetes also raised study limitations including the relatively small amount of data available for people with type 1 diabetes in these primary care electronic medical records, the need to use an algorithm to estimate diabetes type because of the lack of specificity about this in Canadian electronic medical records, the lack of data about education level to better identify the contribution of income and education to glycemic control, and the lack of data about race and ethnicity, which strongly impact outcomes for people with diabetes in Canada and elsewhere. Team members with diabetes also questioned whether HbA1c results might have differed during the 10-year span of the study given the introduction of new technologies in Canada between 2010 and 2019 that allow greater glycemic control.

With respect to implications for policy and future research, people living with diabetes noted that an HbA1c level under or equal to 7.0% appears to be very difficult to reach and maintain for many people living with diabetes in Canada. They suggested that these results be communicated to health professionals to help set realistic expectations, and also suggested that people living with diabetes should consider being “insistent” with their health professionals to explore options for treatment and appropriate goals. Although guidelines suggest that HbA1c targets should be set between a health professional and a person living with diabetes while taking into account all relevant aspects of the person’s life, in practice, many people with diabetes do not receive this level of individualized care. [24]

Indigenous patient partners expressed interest in data specifically for Indigenous peoples in Canada. The National Diabetes Repository does not currently contain data from health centres specifically serving Indigenous communities, though there may be data from urban Indigenous peoples within the dataset that cannot be identified separately from the larger data set. Our goal with this project was to establish a means for people living with diabetes to determine research questions and drive epidemiological cohort studies. Although we knew our data source would not allow us to answer research questions specific to Indigenous peoples, we specifically included Indigenous patient partners, researchers, and non-Indigenous researchers who work with Indigenous communities in the project to help us collectively ensure that our approaches would not harm potential future Indigenous-led projects conducted under relevant ethical frameworks such as the First Nations’ Principles of Ownership, Control, Access, and Possession. [57]

## Discussion

In this study codesigned with people living with diabetes, we aimed to explore differences in HbA1c between people with type 1 or type 2 diabetes of different sexes at different ages and with different levels of socioeconomic status, using a large database of primary care electronic medical records for people with diabetes in Canada. We report six main observations from our study.

First, the differences in statistical significance between the smaller sample of people with type 1 diabetes and the much larger sample of people with type 2 diabetes are reflective of broader patterns in research. These patterns have policy implications that can affect the lives of people living with more or less common conditions, including more or less common types of diabetes. Larger sample sizes allow identification of smaller associations or effects. [58] In the case of diabetes, this means that it is easier to identify statistically-significant effects in the much larger populations of people with type 2 diabetes than in the smaller populations of people with type 1 diabetes or the even smaller populations of people with other types. This can have negative policy impacts on people with diabetes, for different reasons. For people with type 2 diabetes, as health research enters the era of big data and personalized medicine, large datasets analyzed by research teams with little clinical, epidemiological, or personal expertise may allow identification of associations or effects that may not be clinically or personally meaningful. For people with type 1 diabetes, who represent an estimated 5-10% of cases of diabetes, minority status within the larger disease community has led to policy issues such as type 2 diabetes being identified as a risk factor for severe COVID-19 outcomes while type 1 diabetes, which demonstrated higher odds ratios or hazard ratios for severe COVID-19 outcomes in multiple studies but had wider confidence intervals due to smaller populations [59–61], was identified only as a “potential risk factor.” [62] As with other less common conditions, it is important that policy decisions that affect people with diabetes account for differences in type of diabetes and avoid applying the same statistical standards to differently-sized populations without accounting for the influence of sample size.

Second, age is an important consideration for HbA1c targets for people with both types of diabetes. Like studies in other countries [7–10,63,64], our study demonstrated overall higher HbA1c values among adolescents with diabetes compared to people with diabetes in other age groups. This may reflect the ways in which adolescents differ from people in other age groups due to biology (e.g., puberty, menarche), diabetes management (e.g., time since diagnosis to develop useful habits and patterns, lack of full control over management options due to family preferences and finances), and life stage (e.g., externally-imposed structures of school, work, family; internally-directed focus on social development.) Although providing high quality health care to children and adolescents with diabetes has long been an area of focus in Canada, [65] our results suggest that adolescents may still need more support both within and outside of clinical encounters.

Third, there is a tendency toward different patterns across the lifecourse between people of different sexes with type 1 diabetes. Our relatively small sample of people with type 1 diabetes in this dataset does not allow us to draw definitive conclusions. However, women living with type 1 diabetes on our team noted that the potential pattern we observed mimics their own HbA1c patterns during childbearing, perimenopausal, and menopausal years. Hermann and colleagues similarly observed significantly higher HbA1c among female people with type 1 diabetes compared to male people prior to the age of 30 years and after the age of 50 years. [66] People with type 1 diabetes who plan to bear children may be particularly motivated to maintain a lower HbA1c during their childbearing years due to more stringent recommendations regarding glycemic control during pregnancy [67,68] and societal, medical, and self-directed expectations regarding how pregnant people, especially those at increased risk, should prioritize the health of their offspring. [69–72] Following childbearing, the hormonal shifts of perimenopause and menopause combined with common life stressors of middle age and gendered parenting roles may explain somewhat higher HbA1c values among many middle-aged female people. As noted by the lead patient partner in this project (DG), the evidence available about menopause and type 1 diabetes is scarce, with a few studies about age of menopause [73–76] and associated health risks [77], but very little evidence about how to manage one’s diabetes and other health concerns post-menopause. [78] This evidence gap impacts negatively on the lives of people with the condition who progress through menopause.

Fourth, the direction of the overall sex-based difference we observed among people with type 2 diabetes differed from some previous studies in other countries. In our study using data from people in Canada, male people with type 2 diabetes had higher overall HbA1c values, indicating potentially higher risk of diabetes-related complications compared to female people. Other studies using data from people in Portugal [13], Brazil and Venezuela [15], Korea [19], and Spain [79] reported the opposite, with overall higher HbA1c values among those who are female compared to those who are male. Studies in the USA [22] and the Netherlands [23] reported no sex-based differences for HbA1c values and a study in Sweden reported higher HbA1c values for male people compared to females. [9] The lack of agreement among different studies regarding sex-based differences might occur because overall differences may be a product of both biology and gender equality as seen via social roles. We note that the World Economic Forum’s Global Gender Gap Report offers data in support of the suggestion that gender roles may explain the different results regarding sex-based differences across countries. As measured by this index, the countries in which male people have lower HbA1c values than female people (Spain, Korea, Portugal, Brazil, Venezuela) have lower mean gender equality (mean 0.723, SD 0.043) than the countries in which there was no difference (USA, Netherlands: mean 0.763, SD 0.001), which in turn had lower mean gender equality than countries in which male people had higher HbA1c values (Canada, Sweden: mean 0.798, SD 0.036). [80] In other words, in countries with better overall gender equality, women with type 2 diabetes may have better health relative to men than women with type 2 diabetes in countries with worse gender equality.

Fifth, people with type 2 diabetes living in less affluent areas in Canada had higher HbA1c values than those living in more affluent areas. This is unfortunately unsurprising, as type 2 diabetes is a progressive disease and is more prevalent in Canada among people with lower incomes than among those with higher incomes. [81–83] There may also be a similar pattern among people with type 1 diabetes that is not detectable in our relatively small sample. People with type 1 diabetes with higher incomes have less variability in their HbA1c results [12], lower income has been shown to be associated with higher HbA1c among children with type 1 diabetes in Canada [84], and complications are more prevalent among people living with type 1 diabetes in Canada with lower incomes. [85] Because of uneven coverage of diabetes medications and devices across Canada, unequal access to high quality health care, and large differences in levels of food security, people with both types of diabetes who live on lower incomes may face additional challenges in diabetes management compared to those with higher incomes. [86–89] Without effort to address these inequities, such patterns may worsen in the coming years with the advent of new technologies and medications.

Sixth and finally, substantial proportions of people with diabetes in Canada are not yet demonstrating guideline-recommended HbA1c values. This shows the difficulty of reaching and maintaining this goal. [90] Aronson and colleagues similarly showed within a larger sample of 3600 adults living with type 1 diabetes and receiving care from endocrinologists in Canada that less than a quarter of people were achieving HbA1c ≤7.0%. [91] Health professionals and policymakers should be aware of this gap to better support people living with diabetes in Canada. As noted by people on our team who live with diabetes, health professionals’ acknowledgement of the difficulty of attaining this target would help them feel less like they are failing, and more like they are part of a large group living with a difficult condition and potentially struggling to achieve targets set by academic researchers and health professionals. Policymakers can improve this situation by better funding health care, education, medications (e.g., insulin, other medications), supplies (e.g., test strips, flash or continuous glucose monitors, closed loop artificial pancreas systems), [92] food security initiatives (e.g., access to affordable healthy foods), [93] healthy environment initiatives (e.g., walking trails, bicycle paths, community gardens), [94] broader anti-poverty initiatives, [95] and research aimed at supporting people living with diabetes in Canada in achieving self-directed health goals. [96]

This study has three main limitations. First, although the data set was large and of overall high quality, the lack of relevant data (e.g., electronic medical records in Canada do not include relevant data such as ethnicity), the use of proxy variables (e.g., socioeconomic status as a quintile derived from postal codes) and the necessity of using an algorithm to predict type of diabetes may have limited our results. We cannot be certain that all people predicted by the algorithm to have type 1 diabetes were correctly identified, and even then, the small number of people identified by the algorithm as having type 1 diabetes mean that our findings with respect to type 1 diabetes are less conclusive than those for type 2 diabetes. Other, rarer types of diabetes such as latent autoimmune diabetes in adults (LADA) are not represented at all. Second, we did not include comorbid medical conditions in this preliminary study, as these were not part of the research question identified by people living with diabetes. Although people who live with other conditions in addition to diabetes may have higher or lower HbA1c values than those who live only with diabetes, for this preliminary analysis, we sought only to determine broad patterns in this large, national dataset. Third, our analyses had some threats to external validity (i.e., generalizability). All the medical records from the National Diabetes Repository were collected from primary care records from 5 provinces in Canada. Approximately 15% of people in Canada lack access to a primary health care provider in Canada, and lack of access is not distributed evenly [97,98], meaning that our study may have some selection bias.

This study also has three main strengths. First, the entire study, from the initial idea and development of the research question to interpretation of results and drafting of this manuscript, was done in full partnership with people living with the condition studied. This allowed us to identify a research question that is relevant to people living with diabetes in Canada, enrich our interpretation of results, and avoid framing our results in ways that are heedless of the humanity of people whose medical data was analyzed. Second, this study offers insight about glycemic control for people living with diabetes across Canada, includes key variables of sex and age, and accounts for the potential influence of socioeconomic status. It is important to avoid a one-size-fits-all approach to discussions about diabetes management. Identifying patterns according to common variables is a step towards more individualized care. Third, data from the Canadian National Diabetes Repository represent high-quality big data across multiple provinces, allowing national-level analyses that were previously difficult to perform in Canada.

## Conclusion

This study demonstrates the value and potential of patient-led research and of a national data repository for diabetes. People who live with a condition should have power over setting health research agendas so that research serves their needs. Responding to a research question developed by people living with diabetes, we mapped nearly a million HbA1c results over ten years from people with type 1 and type 2 diabetes in Canada and showed how HbA1c results may differ by age, sex, and socioeconomic status. These factors are important to consider when setting HbA1c targets and when studying relationships between HbA1c and complications of diabetes. Further research and support are needed to help people manage diabetes across life stages, with notable challenges during adolescence for people of both sexes and during menopause for people of female sex. As noted by members of our research team living with diabetes, health professionals should be aware of the difficulty of maintaining a HbA1c value below guideline-recommended targets, and policymakers should provide support for people with diabetes in Canada to live healthy lives.

## Data Availability

National Diabetes Repository data are available for analysis: https://repository.diabetesaction.ca/request-access/

## DECLARATIONS

### Abbreviations

None

### Ethics Approval, Consent to Participate and Consent for Publication

Our study received ethical approval from the Laval University Research Ethics Committee, 2020-373/20-01-2021. Original collection of data was also conducted under ethics approval. This study received approval by the National Diabetes Repository governance committee. The governance committee is made up of minimum 50% people living with diabetes.

## Acknowledgements

The authors gratefully acknowledge the contributions of all team members who contributed greatly to this research project as a whole but did not accept the invitation to participate on this particular paper as co-authors, and also Conrad Pow and Tao Chen for their assistance with the Canadian National Diabetes Repository, and Anne-Sophie Julien for her advice on statistical analyses.

## Supplementary materials

Figure S1 and Table S1 show pairwise interaction contrasts for both types of diabetes. For people with type 1 diabetes, no differences between men and women are statistically significant in these analyses. For people with type 2 diabetes, there are statistically significant sex differences with p-values of <.0001 in nearly all age categories.

**Figure S1.**
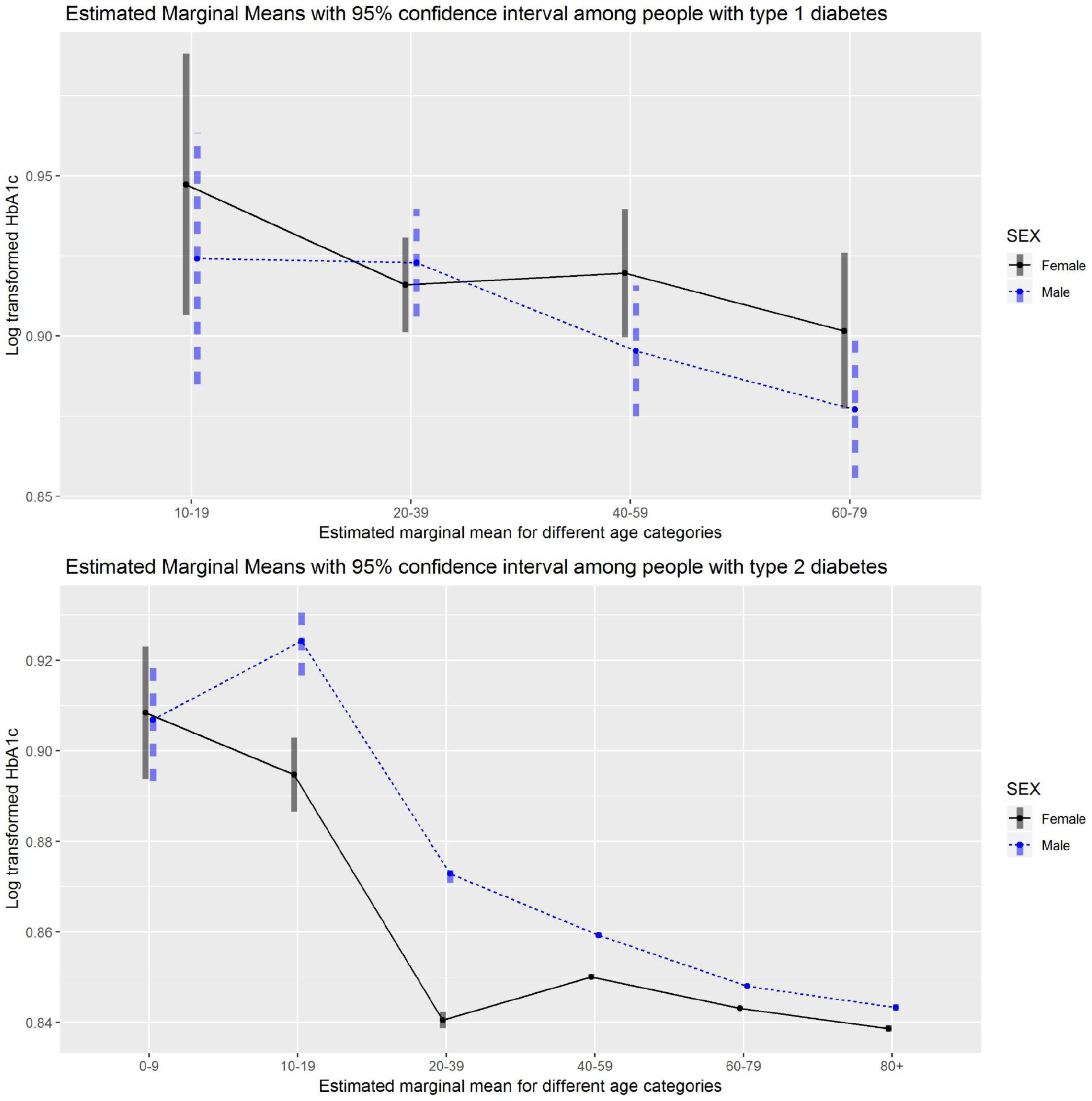
Pairwise contrast for sex differences in HbA1c results in each age categories.

**Table S1.** Pairwise contrast for sex differences in HbA1c results in people with type 1 and 2 diabetes in each age categories.

|  | Type 1 diabetes (total n = 1949 HbA1c results from 296 people) |  |  | Type 2 diabetes (total n = 945,262 HbA1c results from 90,417 people) |  |  |
| --- | --- | --- | --- | --- | --- | --- |
| Age Category | Estimate | Standard Error | P-value | Estimate | Standard Error | P-value |
| 10 - 19 | 0.02314 | 0.0291 | 0.4260 | -0.0295 | 0.00568 | <.0001 |
| 20 - 39 | -0.00689 | 0.0114 | 0.5450 | -0.0324 | 0.00137 | <.0001 |
| 40 - 59 | 0.02426 | 0.0146 | 0.097 | -0.0092 | 0.00043 | <.0001 |
| 60 - 79 | 0.02456 | 0.0165 | 0.1380 | -0.0049 | 0.00027 | <.0001 |
| 80+ | - | - | - | -0.0046 | 0.0005 | <.0001 |

## References

1. Diabetes [Internet]. [cited 2020 Jun 9]. Available from: https://www.who.int/health-topics/diabetes

2. Xu G, Liu B, Sun Y, Du Y, Snetselaar LG, Hu FB, Bao W. Prevalence of diagnosed type 1 and type 2 diabetes among US adults in 2016 and 2017: population based study. BMJ 2018 Sep 4;362:k1497. PMID:30181166

3. Carstensen B, Rønn PF, Jørgensen MM. Prevalence, incidence and mortality of type 1 and type 2 diabetes in Denmark 1996–2016 [Internet]. BMJ Open Diabetes Research & Care. 2020. p. e001071. [doi: 10.1136/bmjdrc-2019-001071]

4. Welsh KJ, Kirkman MS, Sacks DD. Role of Glycated Proteins in the Diagnosis and Management of Diabetes: Research Gaps and Future Directions. Diabetes Care 2016 Aug;39(8):1299–1306. PMID:27457632

5. Silverman RA, Thakker U, Ellman T, Wong I, Smith K, Ito K, Graff K. Hemoglobin A1c as a screen for previously undiagnosed prediabetes and diabetes in an acute-care setting. Diabetes Care Am Diabetes Assoc; 2011 Sep;34(9):1908–1912. PMID:21775751

6. Burson R, Moran KK. Individualizing Targets for Glycemic Control. Home Healthc Now 2018;36(3):190–191. PMID:29722710

7. Pinhas-Hamiel O, Hamiel U, Boyko V, Graph-Barel C, Reichman B, Lerner-Geva L. Trajectories of HbA1c levels in children and youth with type 1 diabetes. PLoS One 2014 Oct 2;9(10):e109109. PMID:25275650

8. Clements MA, Foster NC, Maahs DM, Schatz DA, Olson BA, Tsalikian E, Lee JM, Burt-Solorzano CM, Tamborlane WV, Chen V, Miller KM, Beck RW, T1D Exchange Clinic Network. Hemoglobin A1c (HbA1c) changes over time among adolescent and young adult participants in the T1D exchange clinic registry. Pediatr Diabetes 2016 Aug;17(5):327–336. PMID:26153338

9. Julin B, Willers C, Leksell J, Lindgren P, Looström Muth K, Svensson A-M, Lilja M, Dahlström T. Association between sociodemographic determinants and health outcomes in individuals with type 2 diabetes in Sweden. Diabetes Metab Res Rev 2018 May;34(4):e2984. PMID:29377503

10. Hessler DM, Fisher L, Mullan JT, Glasgow RE, Masharani U. Patient age: a neglected factor when considering disease management in adults with type 2 diabetes. Patient Educ Couns 2011 Nov;85(2):154–159. PMID:21112720

11. Whyte MB, Hinton W, McGovern A, van Vlymen J, Ferreira F, Calderara S, Mount J, Munro N, de Lusignan S. Disparities in glycaemic control, monitoring, and treatment of type 2 diabetes in England: A retrospective cohort analysis. PLoS Med 2019 Oct;16(10):e1002942. PMID:31589609

12. Auzanneau M, Lanzinger S, Bohn B, Kroschwald P, Kuhnle-Krahl U, Holterhus PM, Placzek K, Hamann J, Bachran R, Rosenbauer J, Maier W, DPV Initiative. Area Deprivation and Regional Disparities in Treatment and Outcome Quality of 29,284 Pediatric Patients With Type 1 Diabetes in Germany: A Cross-sectional Multicenter DPV Analysis. Diabetes Care 2018 Dec;41(12):2517–2525. PMID:30327359

13. Góis C, Duarte TA, Paulino S, Raposo JF, do Carmo I, Barbosa A. Depressive symptoms are associated with poor glycemic control among women with type 2 diabetes mellitus. BMC Res Notes 2018 Jan 16;11(1):38. PMID:29338774

14. Willers C, Iderberg H, Axelsen M, Dahlström T, Julin B, Leksell J, Lindberg A, Lindgren P, Looström Muth K, Svensson A-M, Lilja M. Sociodemographic determinants and health outcome variation in individuals with type 1 diabetes mellitus: A register-based study. PLoS One 2018 Jun 29;13(6):e0199170. PMID:29958293

15. G Duarte F, da Silva Moreira S, Almeida M da CC, de Souza Teles CA, Andrade CS, Reingold AL, Moreira ED Jr. Sex differences and correlates of poor glycaemic control in type 2 diabetes: a cross-sectional study in Brazil and Venezuela. BMJ Open 2019 Mar 5;9(3):e023401. PMID:30842107

16. Larkin ME, Backlund J-Y, Cleary P, Bayless M, Schaefer B, Canady J, Nathan DM, Diabetes Control and Complications Trial/Epidemiology of Diabetes Interventions and Complications (DCCT/EDIC) Research Group. Disparity in management of diabetes and coronary heart disease risk factors by sex in DCCT/EDIC. Diabet Med 2010 Apr;27(4):451– 458. PMID:20536518

17. Mair C, Wulaningsih W, Jeyam A, McGurnaghan S, Blackbourn L, Kennon B, Leese G, Lindsay R, McCrimmon RJ, McKnight J, Petrie JR, Sattar N, Wild SH, Conway N, Craigie I, Robertson K, Bath L, McKeigue PM, Colhoun HM, Scottish Diabetes Research Network (SDRN) Epidemiology Group. Glycaemic control trends in people with type 1 diabetes in Scotland 2004-2016. Diabetologia 2019 Aug;62(8):1375–1384. PMID:31104095

18. Kautzky-Willer A, Kosi L, Lin J, Mihaljevic R. Gender-based differences in glycaemic control and hypoglycaemia prevalence in patients with type 2 diabetes: results from patient-level pooled data of six randomized controlled trials. Diabetes Obes Metab 2015 Jun;17(6):533– 540. PMID:25678212

19. Choe S-A, Kim JY, Ro YS, Cho S-I. Women are less likely than men to achieve optimal glycemic control after 1 year of treatment: A multi-level analysis of a Korean primary care cohort. PLoS One 2018 May 2;13(5):e0196719. PMID:29718952

20. Clemens KK, Woodward M, Neal B, Zinman B. Sex Disparities in Cardiovascular Outcome Trials of Populations With Diabetes: A Systematic Review and Meta-analysis. Diabetes Care 2020 May;43(5):1157–1163. PMID:32312859

21. Shah VN, Wu M, Polsky S, Snell-Bergeon JK, Sherr JL, Cengiz E, DiMeglio LA, Pop-Busui R, Mizokami-Stout K, Foster NC, Beck RW, for T1D Exchange Clinic Registry. Gender differences in diabetes self-care in adults with type 1 diabetes: Findings from the T1D Exchange clinic registry. J Diabetes Complications 2018 Oct;32(10):961–965. PMID:30121205

22. Misra R, Lager J. Ethnic and gender differences in psychosocial factors, glycemic control, and quality of life among adult type 2 diabetic patients. J Diabetes Complications 2009 Jan;23(1):54–64. PMID:18413181

23. de Jong M, Vos RC, de Ritter R, van der Kallen CJ, Sep SJ, Woodward M, Stehouwer CDA, Bots ML, Peters SS. Sex differences in cardiovascular risk management for people with diabetes in primary care: a cross-sectional study. BJGP Open [Internet] 2019 Jul;3(2). PMID:31366676

24. Coons MJ, Greiver M, Aliarzadeh B, Meaney C, Moineddin R, Williamson T, Queenan J, Yu CH, White DG, Kiran T, Kane JJ. Is glycemia control in Canadians with diabetes individualized? A cross-sectional observational study. BMJ Open Diabetes Res Care 2017 Jun 8;5(1):e000316. PMID:28761645

25. Government of Canada, Canadian Institutes of Health Research, Research, Translation K. Strategy for Patient-Oriented Research - CIHR [Internet]. 2018 [cited 2020 Jun 9]. Available from: https://cihr-irsc.gc.ca/e/41204.html

26. Witteman HO, Chipenda Dansokho S, Colquhoun H, Fagerlin A, Giguere AMC, Glouberman S, Haslett L, Hoffman A, Ivers NM, Légaré F, Légaré J, Levin CA, Lopez K, Montori VM, Renaud J-S, Sparling K, Stacey D, Volk RR. Twelve Lessons Learned for Effective Research Partnerships Between Patients, Caregivers, Clinicians, Academic Researchers, and Other Stakeholders. J Gen Intern Med 2018 Apr;33(4):558–562. PMID:29327211

27. Dogba MJ, Dipankui MT, Chipenda Dansokho S, Légaré F, Witteman HH. Diabetes-related complications: Which research topics matter to diverse patients and caregivers? Health Expect 2018 Apr;21(2):549–559. PMID:29165920

28. Diabetes Action Canada - SPOR network [Internet]. 2016 [cited 2021 Feb 12]. Available from: https://repository.diabetesaction.ca/

29. Willison DJ, Trowbridge J, Greiver M, Keshavjee K, Mumford D, Sullivan F. Participatory governance over research in an academic research network: the case of Diabetes Action Canada. BMJ Open 2019 Apr 20;9(4):e026828. PMID:31005936

30. Weisman A, Tu K, Young J, Kumar M, Austin PC, Jaakkimainen L, Lipscombe L, Aronson R, Booth GG. Validation of a type 1 diabetes algorithm using electronic medical records and administrative healthcare data to study the population incidence and prevalence of type 1 diabetes in Ontario, Canada. BMJ Open Diabetes Res Care [Internet] 2020 Jun;8(1). PMID:32565422

31. cphi-toolkit-area-level-measurement-pccf-2018-en-web.pdf. Available from: https://www.cihi.ca/sites/default/files/document/cphi-toolkit-area-level-measurement-pccf-2018-en-web.pdf

32. [No title] [Internet]. [cited 2021 Jul 1]. Available from: https://repository.diabetesaction.ca/wp-content/uploads/2020/11/ndr_ses.html

33. Pekár S, Brabec M. Generalized estimating equations: A pragmatic and flexible approach to the marginal GLM modelling of correlated data in the behavioural sciences. Ethology Wiley; 2018 Feb;124(2):86–93.

34. Curran-Everett D. Explorations in statistics: the log transformation. Adv Physiol Educ 2018 Jun 1;42(2):343–347. PMID:29761718

35. Schweiger B, Klingensmith GJ, Snell-Bergeon JJ. Menarchal timing in type 1 diabetes through the last 4 decades. Diabetes Care 2010 Dec;33(12):2521–2523. PMID:20843975

36. Raha O, Sarkar B, Godi S, GhoshRoy A, Pasumarthy V, Chowdhury S, JDRF-India, Vadlamudi RR. Menarcheal age of type 1 diabetic Bengali Indian females. Gynecol Endocrinol 2013 Nov;29(11):963–966. PMID:23952104

37. Mukherjee MS, Coppenrath VA, Dallinga BB. Pharmacologic management of types 1 and 2 diabetes mellitus and their complications in women of childbearing age. Pharmacotherapy 2015 Feb;35(2):158–174. PMID:25644781

38. Rasmussen B, Nankervis A, Skouteris H, McNamara C, Nagle C, Steele C, Bruce L, Holton S, Wynter K. Factors associated with breastfeeding to 3 months postpartum among women with type 1 and type 2 diabetes mellitus: An exploratory study. Women Birth 2020 May;33(3):e274.#x2013;e279. PMID:31239238

39. Gaudio M, Dozio N, Feher M, Scavini M, Caretto A, Joy M, Van Vlymer J, Hinton W, de Lusignan S. Trends in Factors Affecting Pregnancy Outcomes Among Women With Type 1 or Type 2 Diabetes of Childbearing Age (2004-2017). Front Endocrinol 2020;11:596633. PMID:33692751

40. Caruso S, Cianci A, Cianci S, Monaco C, Fava V, Cavallari V. Ultrastructural Study of Clitoral Cavernous Tissue and Clitoral Blood Flow From Type 1 Diabetic Premenopausal Women on Phosphodiesterase-5 Inhibitor. J Sex Med 2019 Mar;16(3):375–382. PMID:30773497

41. Purnamasari D, Puspitasari MD, Setiyohadi B, Nugroho P, Isbagio H. Low bone turnover in premenopausal women with type 2 diabetes mellitus as an early process of diabetes-associated bone alterations: a cross-sectional study [Internet]. BMC Endocrine Disorders. 2017. [doi: 10.1186/s12902-017-0224-0]

42. Sjöberg L, Pitkäniemi J, Harjutsalo V, Haapala L, Tiitinen A, Tuomilehto J, Kaaja R. Menopause in women with type 1 diabetes. Menopause 2011 Feb;18(2):158–163. PMID:20881890

43. Khalil N, Sutton-Tyrrell K, Strotmeyer ES, Greendale GA, Vuga M, Selzer F, Crandall CJ, Cauley JJ. Menopausal bone changes and incident fractures in diabetic women: a cohort study. Osteoporos Int 2011 May;22(5):1367–1376. PMID:20658126

44. Scott AR, Dhindsa P, Forsyth J, Mansell P, Kliofem Study Collaborative Group. Effect of hormone replacement therapy on cardiovascular risk factors in postmenopausal women with diabetes. Diabetes Obes Metab 2004 Jan;6(1):16–22. PMID:14686958

45. Wickham H. Easily Install and Load the “Tidyverse” [R package tidyverse version 1.2.1]. Comprehensive R Archive Network (CRAN); 2017 [cited 2021 Jul 8]; Available from: https://CRAN.R-project.org/package=tidyverse

46. Hadley Wickham RF, Henry L, Müller K. dplyr: A Grammar of Data Manipulation, 2019. R package version 0801 [Internet] 2019;3. Available from: https://CRAN.R-project.org/package=dplyr

47. Wickham H. The Split-Apply-Combine Strategy for Data Analysis. Journal of Statistical Software, Articles 2011;40(1):1–29.

48. Robinson D, Hayes A. broom: Convert statistical analysis objects into tidy tibbles. Version 0 5 2 2019;

49. Dowle M, Srinivasan A, Short T, Lianoglou S. data. table: Extension of “data. frame.” R package version 1122 2019;1(8).

50. MuMIn BB. Multi-model inference. R package version 1.43. 17. 2020. 2020.

51. Fox J, Weisberg S. An R Companion to Applied Regression. SAGE Publications; 2019. ISBN:9781544336480

52. Højsgaard S, Halekoh U, Yan J. The R Package geepack for Generalized Estimating Equations. Journal of Statistical Software, Articles 2006;15(2):1–11.

53. Yan J, Fine J. Estimating equations for association structures. Stat Med 2004 Mar 30;23(6):859–74; discussion 875–7,879–80. PMID:15027075

54. Yan J. Geepack: yet another package for generalized estimating equations. R-news 2002;2(3):12–14.

55. Lenth RR. Estimated marginal means, aka least-squares means [R Package Emmeans Version 1.6. 1]. 2021.

56. Radin MM. Pitfalls in hemoglobin A1c measurement: when results may be misleading. J Gen Intern Med 2014 Feb;29(2):388–394. PMID:24002631

57. The First Nations Principles of OCAP® [Internet]. 2020 [cited 2021 Jun 18]. Available from: https://fnigc.ca/ocap-training/

58. Groft SC, Paz MM. Rare diseases--avoiding misperceptions and establishing realities: the need for reliable epidemiological data. Rare Diseases Epidemiology Springer Netherlands; 2010;3–14.

59. Gregory JM, Slaughter JC, Duffus SH, Smith TJ, LeStourgeon LM, Jaser SS, McCoy AB, Luther JM, Giovannetti ER, Boeder S, Pettus JH, Moore DD. COVID-19 Severity Is Tripled in the Diabetes Community: A Prospective Analysis of the Pandemic’s Impact in Type 1 and Type 2 Diabetes. Diabetes Care 2021 Feb;44(2):526–532. PMID:33268335

60. Barron E, Bakhai C, Kar P, Weaver A, Bradley D, Ismail H, Knighton P, Holman N, Khunti K, Sattar N, Wareham NJ, Young B, Valabhji J. Associations of type 1 and type 2 diabetes with COVID-19-related mortality in England: a whole-population study. Lancet Diabetes Endocrinol 2020 Oct;8(10):813–822. PMID:32798472

61. McGurnaghan SJ, Weir A, Bishop J, Kennedy S, Blackbourn LAK, McAllister DA, Hutchinson S, Caparrotta TM, Mellor J, Jeyam A, O’Reilly JE, Wild SH, Hatam S, Höhn A, Colombo M, Robertson C, Lone N, Murray J, Butterly E, Petrie J, Kennon B, McCrimmon R, Lindsay R, Pearson E, Sattar N, McKnight J, Philip S, Collier A, McMenamin J, Smith-Palmer A, Goldberg D, McKeigue PM, Colhoun HM, Public Health Scotland COVID-19 Health Protection Study Group, Scottish Diabetes Research Network Epidemiology Group. Risks of and risk factors for COVID-19 disease in people with diabetes: a cohort study of the total population of Scotland. Lancet Diabetes Endocrinol 2021 Feb;9(2):82–93. PMID:33357491

62. Cooney E. For people with type 1 diabetes, CDC Covid guidelines are puzzling. STAT [Internet] 2021 Jan 11 [cited 2021 Jul 21]; Available from: https://www.statnews.com/2021/01/11/for-people-with-type-1-diabetes-cdc-guidelines-for-covid-19-vaccine-priority-are-puzzling/comment-page-1/

63. Foster NC, Beck RW, Miller KM, Clements MA, Rickels MR, DiMeglio LA, Maahs DM, Tamborlane WV, Bergenstal R, Smith E, Olson BA, Garg SS. State of Type 1 Diabetes Management and Outcomes from the T1D Exchange in 2016-2018. Diabetes Technol Ther 2019 Feb;21(2):66–72. PMID:30657336

64. Beck RW, Tamborlane WV, Bergenstal RM, Miller KM, DuBose SN, Hall CA, T1D Exchange Clinic Network. The T1D Exchange clinic registry. J Clin Endocrinol Metab 2012 Dec;97(12):4383–4389. PMID:22996145

65. Canadian Diabetes Association. Clinical Practice Guidelines Expert Committee. Canadian Diabetes Association 2013 clinical practice guidelines for the prevention and management of diabetes in Canada. Canadian Diabetes Association; 2013.

66. Hermann JM, Miller KM, Hofer SE, Clements MA, Karges W, Foster NC, Fröhlich-Reiterer E, Rickels MR, Rosenbauer J, DeSalvo DJ, Holl RW, Maahs DM, T1D Exchange Clinic Network and the DPV initiative. The Transatlantic HbA1c gap: differences in glycaemic control across the lifespan between people included in the US T1D Exchange Registry and those included in the German/Austrian DPV registry. Diabet Med 2020 May;37(5):848–855. PMID:31557351

67. Diabetes and Pregnancy [Internet]. [cited 2021 Jul 16]. Available from: https://guidelines.diabetes.ca/cpg/chapter36

68. Murphy HH. Continuous glucose monitoring targets in type 1 diabetes pregnancy: every 5% time in range matters. Diabetologia 2019 Jul;62(7):1123–1128. PMID:31161344

69. Myers ST, Grasmick HH. The Social Rights and Responsibilities of Pregnant Women: An Application of Parsons’s Sick Role Model. J Appl Behav Sci SAGE Publications Inc; 1990 May 1;26(2):157–172.

70. Berg M, Honkasalo MM. Pregnancy and diabetes--a hermeneutic phenomenological study of women’s experiences. J Psychosom Obstet Gynaecol 2000 Mar;21(1):39–48. PMID:10907214

71. Berg M, Sparud-Lundin C. Experiences of professional support during pregnancy and childbirth - a qualitative study of women with type 1 diabetes. BMC Pregnancy Childbirth 2009 Jul 3;9:27. PMID:19575789

72. McGrath M, Chrisler JJ. A lot of hard work, but doable: Pregnancy experiences of women with type-1 diabetes. Health Care Women Int 2017 Jun;38(6):571–592. PMID:27918866

73. Yarde F, van der Schouw YT, de Valk HW, Franx A, Eijkemans MJC, Spiering W, Broekmans FJM, OVADIA study group. Age at menopause in women with type 1 diabetes mellitus: the OVADIA study. Hum Reprod 2015 Feb;30(2):441–446. PMID:25452435

74. Yi Y, El Khoudary SR, Buchanich JM, Miller RG, Rubinstein D, Matthews K, Orchard TJ, Costacou T. Women with Type 1 diabetes (T1D) experience a shorter reproductive period compared with nondiabetic women: the Pittsburgh Epidemiology of Diabetes Complications (EDC) study and the Study of Women’s Health Across the Nation (SWAN). Menopause [Internet] LWW; 2021; Available from: https://journals.lww.com/menopausejournal/Abstract/9000/Women_with_Type_1_diabetes__T1D__experience_a.96999.aspx

75. Yi Y, El Khoudary SR, Buchanich JM, Miller RG, Rubinstein D, Orchard TJ, Costacou T. Association of age at diabetes complication diagnosis with age at natural menopause in women with type 1 diabetes: The Pittsburgh Epidemiology of Diabetes Complications (EDC) Study. J Diabetes Complications 2021 Mar;35(3):107832. PMID:33446412

76. Yi Y, El Khoudary SR, Buchanich JM, Miller RG, Rubinstein D, Orchard TJ, Costacou T. Predictors of the age at which natural menopause occurs in women with type 1 diabetes: the Pittsburgh Epidemiology of Diabetes Complications (EDC) study. Menopause 2021 Apr 5;28(7):735–740. PMID:33828035

77. Keshawarz A, Pyle L, Alman A, Sassano C, Westfeldt E, Sippl R, Snell-Bergeon J. Type 1 Diabetes Accelerates Progression of Coronary Artery Calcium Over the Menopausal Transition: The CACTI Study. Diabetes Care 2019 Dec;42(12):2315–2321. PMID:31558547

78. Mackay L, Kilbride L, Adamson KA, Chisholm J. Hormone replacement therapy for women with type 1 diabetes mellitus. Cochrane Database Syst Rev 2013 Jun 6;(6):CD008613. PMID:23744560

79. Galbete A, Cambra K, Forga L, Baquedano FJ, Aizpuru F, Lecea O, Librero J, Ibáñez B. Achievement of cardiovascular risk factor targets according to sex and previous history of cardiovascular disease in type 2 diabetes: A population-based study. J Diabetes Complications 2019 Dec;33(12):107445. PMID:31668588

80. World Economic Forum. Global Gender Gap Report 2021: Insight Report [Internet]. World Economic Forum; 2021 Mar. Available from: http://www3.weforum.org/docs/WEF_GGGR_2021.pdf

81. Dinca-Panaitescu M, Dinca-Panaitescu S, Raphael D, Bryant T, Pilkington B, Daiski I. The dynamics of the relationship between diabetes incidence and low income: longitudinal results from Canada’s National Population Health Survey. Maturitas 2012 Jul;72(3):229–235. PMID:22551632

82. Lysy Z, Booth GL, Shah BR, Austin PC, Luo J, Lipscombe LL. The impact of income on the incidence of diabetes: a population-based study. Diabetes Res Clin Pract 2013 Mar;99(3):372–379. PMID:23305902

83. Bird Y, Lemstra M, Rogers M, Moraros J. The relationship between socioeconomic status/income and prevalence of diabetes and associated conditions: A cross-sectional population-based study in Saskatchewan, Canada. Int J Equity Health 2015 Oct 12;14:93. PMID:26458543

84. Zuijdwijk CS, Cuerden M, Mahmud FF. Social determinants of health on glycemic control in pediatric type 1 diabetes. J Pediatr 2013 Apr;162(4):730–735. PMID:23360562

85. Butalia S, Patel AB, Johnson JA, Ghali WA, Rabi DD. Geographic Clustering of Acute Complications and Sociodemographic Factors in Adults with Type 1 Diabetes. Can J Diabetes 2017 Apr;41(2):132–137. PMID:27887926

86. Beryl Pilkington F, Daiski I, Bryant T, Dinca-panaitescu M, Dinca-panaitescu S, Raphael D. The Experience of Living with Diabetes for Low-income Canadians. Canadian Journal of Diabetes 2010 Jan 1;34(2):119–126.

87. Visekruna S, McGillis Hall L, Parry M, Spalding K. Intersecting Health Policy and the Social Determinants of Health in Pediatric Type 1 Diabetes Management and Care. J Pediatr Nurs 2017 Nov;37:62–69. PMID:28683888

88. Campbell RB, Larsen M, DiGiandomenico A, Davidson MA, Booth GL, Hwang SW, McBrien KA, Campbell DJT. The challenges of managing diabetes while homeless: a qualitative study using photovoice methodology. CMAJ CMAJ; 2021 Jul 12;193(27):E1034– E1041.

89. Hershey JA, Morone J, Lipman TH, Hawkes CC. Social Determinants of Health, Goals and Outcomes in High-Risk Children With Type 1 Diabetes. Can J Diabetes 2021 Jul;45(5):444–450.e1. PMID:33863638

90. Ivers NM, Jiang M, Alloo J, Singer A, Ngui D, Casey CG, Yu CC. Diabetes Canada 2018 clinical practice guidelines: Key messages for family physicians caring for patients living with type 2 diabetes. Can Fam Physician 2019 Jan;65(1):14–24. PMID:30674509

91. Aronson R, Brown RE, Abitbol A, Goldenberg R, Yared Z, Ajala B, Yale J-F. The Canadian LMC Diabetes Registry: A Profile of the Demographics, Management, and Outcomes of Individuals with Type 1 Diabetes. Diabetes Technol Ther 2021 Jan;23(1):31–40. PMID:32667835

92. Haque WZ, Demidowich AP, Sidhaye A, Golden SH, Zilbermint M. The Financial Impact of an Inpatient Diabetes Management Service. Curr Diab Rep 2021 Jan 15;21(2):5. PMID:33449246

93. Ippolito MM, Lyles CR, Prendergast K, Marshall MB, Waxman E, Seligman HH. Food insecurity and diabetes self-management among food pantry clients. Public Health Nutr 2017 Jan;20(1):183–189. PMID:27406399

94. Sidawi B, Alhariri MT, Albaker WW. Creating a healthy built environment for diabetic patients: the case study of the eastern province of the Kingdom of Saudi Arabia. Glob J Health Sci 2014 Apr 14;6(4):136–147. PMID:24999135

95. Hsu C-C, Lee C-H, Wahlqvist ML, Huang H-L, Chang H-Y, Chen L, Shih S-F, Shin S-J, Tsai W-C, Chen T, Huang C-T, Cheng J-S. Poverty increases type 2 diabetes incidence and inequality of care despite universal health coverage. Diabetes Care 2012 Nov;35(11):2286–2292. PMID:22912425

96. What we fund [Internet]. [cited 2021 Sep 27]. Available from: https://www.diabetes.ca/research/what-we-fund

97. Glauser W. Primary care system outdated and inconvenient for many millennials. CMAJ 2018 Dec 3;190(48):E1430–E1431. PMID:30510054

98. Statistics Canada. Primary health care providers, 2019 [Internet]. Government of Canada; 2020 Oct. Report No.: 82-625-X. Available from: https://www150.statcan.gc.ca/n1/pub/82-625-x/2020001/article/00004-eng.htm

